# Longitudinal Assessment of Mental Health in Antarctica Expeditioners

**DOI:** 10.1101/2023.12.07.23299643

**Authors:** S Nagendran, Aftab Ahmad, Ravindra Nath, Shalini Nangalia, Chandrakant Maurya, Ganga Dutt D, Aakash Kanojia

## Abstract

Antarc’ca is known for its harsh climate and long, dark winters. Scien’sts and support staff posted in various base sta’ons in Antarc’ca face unique challenges, which include social isola’on, lack of communica’on from the outside world and prolonged working hours. This may lead to changes in their mental health status during their pos’ng. A longitudinal observa’onal study is planned to evaluate the mental health status and changes in it among the Antarc’c expedi’oners. A detailed protocol is presented in this paper.

## Introduction

The Antarctic, with its extreme conditions, poses unique challenges to the mental well-being of expeditioners.^1^ This study aims to thoroughly and over time assess how people cope psychologically with the isolation, harsh weather, and limited support in Antarctica. The goal is to gather insights that can guide specific help and support systems tailored to this extraordinary environment.

Antarctic expeditions come with a kind of isolation that’s not just physical but impacts the mind negatively. The extreme cold, long periods of darkness, and unpredictable weather make it even tougher. Expeditioners have to deal not only with the tough environment but also with being far away from immediate help. Living away from family and friends further complicates the situation.^2,3^ So, it’s crucial to understand how people adapt and handle the unique challenges of living and working in Antarctica.

This study plans to assess the mental health of expeditioners over time. We plan to collect data before, during, and after their time in Antarctica. We’re bringing together participants from different research stations and using various psychological assessments. These tests will cover their mental well-being, stress levels, sleep quality, and how they cope, and how they connect with others in the isolated Antarctic communities.

In addition to the regular psychological assessments, we will also check how well they are coping with the stress of the Antarctic environment. This comprehensive approach is meant to give us a full picture of mental health, helping us understand how people navigate the challenges of living in Antarctica.

After collecting all this data, we will use advanced statistical methods to find patterns and trends. We want to see how different factors, like isolation and extreme weather, relate to mental health. Looking at the dAnnexure ata over time will help us pinpoint when expeditioners might need support the most.

The ultimate goal is to use what we learn to develop practical ways to support the mental well-being of expeditioners. As we continue exploring and working in Antarctica, taking care of the mental health of those on these missions becomes not just a scientific concern but a responsibility we must meet. This study is our way of tackling that responsibility by using rigorous scientific methods to understand and address the unique mental health challenges posed by the Antarctic environment.

## Lacunae in Existing Knowledge

The unique challenges posed by the Antarctic environment, including extreme isolation, harsh weather conditions, and limited external support, can have profound effects on the mental well-being of expeditioners.^4^ Understanding the nuances of mental health in such an extreme setting is crucial for developing targeted interventions and support systems. There is very limited data pertaining to the trends of changes in Mental Health status among Indian Expeditioners.^5^ This study aims to provide a comprehensive and longitudinal assessment of mental health among Antarctica expeditioners to inform strategies for maintaining well-being and optimizing performance.

## Aims & Objectives

### Aim

To assess the mental health of scientists and staff posted in Antarctica expedition from India and study the changes in it in a one-year time.

### Objectives

Primary - To assess the levels of Depression, Anxiety and Stress of Antarctica expeditioners over a one-year period.

Secondary - To identify patterns and changes in Depression, Anxiety and Stress over a course of one year stay with various factors and the coping mechanisms adopted by them.

## Material & Methods

This is an observational longitudinal study which is planned to be conducted in the Bharti and Maitri station of Antarctica, subject to approval of the competent authorities. The tentative dates of study is during the calendar year of 2024, and all scientist and support staff posted in these Bharti and Maitri stations of Antarctica during the study duration. Universal sampling of all scientist and support staff will be done, hence no sample size estimation is needed. The inclusion criteria to oft the study is that the participants should be posted in either of the two Indian stations, viz. Bharti or Maitri during the study duration and should be willing to fill the questionnaire. Any participant having pre-existing mental health condition or on medication will be excluded from the study.

### Study Tools

The DASS 21 is a 21 item self-report questionnaire designed to measure the severity of a range of symptoms common to both Depression and Anxiety. In. completing the DASS, the individual is required to indicate the presence of a symptom over the previous week. Each item is scored from 0 (did not apply to me at all over the last week) to 3 (applied to me very much or most of the time over the past week).

The essential function of the DASS is to assess the severity of the core symptoms of Depression, Anxiety and Stress. Accordingly, the DASS allows not only a way to measure the severity of a patient’s symptoms but a means by which a patient’s response to treatment can also be measured.

The final questionnaire will include socio-demographic profile of the participants along with DASS-21.

### Ethical Considerations

Ethical Clearance for the study is obtained from the Institutional Ethical of Teerthanker Mahaveer Medical College and Research Centre, which is affiliated to Teerthanker Mahaveer University, vide letter number TMU/IECNov23/80 dated 24^th^ November 2023. Written Informed consent will be taken from the Expeditioners & will be kept strictly confidential.

Data collected during the study will be used for academic purposes alone and no personal information of the study subjects will be divulged to anyone.

Any participant who is diagnosed with Depression, Anxiety and Stress will be offered support through teleconsultation from the Department of Psychiatry and Department of Community Medicine, TMMC&RC, Moradabad. Detailed psychological evaluation will be carried out using relevant tools by a senior faculty of a tertiary care hospital. Medicines, if needed, is available at both the base stations.

### Expected Outcomes

Patients diagnosed with constant high levels of Depression, Anxiety and Stress will be assessed with specific questionnaires such as HAM-A, HAM-D and BPRS. These participants will also be offered free counselling & treatment support through teleconsultation from Department of Psychiatry, Teerthanker Mahaveer Medical College & Research Centre, TMU Moradabad.

## Conclusion

The present study is conceptualized to assess the mental health of expedi’oners posted in Antarc’ca and will provide valuable insight into the changes seen in mental health status of the Antarc’c expedi’oners during their course of stay, as well as the coping strategies adopted by them. This will help strengthen the pre-pos’ng psychological evalua’on of Antarc’c expedi’oners.

## Supporting information

Institutional Ethical Committee Approval Letter

## Data Availability

Data collection is yet to commence. Data once collected will be available upon reasonable request to the authors.

