## Supplementary material for "Longitudinal Assessment of Mental Health in Antarctica Expeditioners": Institutional Ethical Committee Approval Letter

Accredited with NAAC A Grade
12-B Status from UGC

### TEERTHANKER MAHAVEER UNIVERSITY

(Established under Govt. of U. P. Act No. 30, 2008)

**Delhi Road, Moradabad (U.P)**

**INSTITUTIONAL ETHICS COMMITTEE**

**Ref. No.** TMU/IEC Nov. 23/80 **Dated :** 24/11/2023

#### ETHICAL CLEARANCE CERTIFICATE

**PROJECT TITLE: Longitudinal Assessment of Mental Health in Antarctica Expeditioners**

**STUDY TEAM: Dr. Ravindra Nath, Dr. S Nagendran, Dr. Aftab Ahmad**

**DEPARTMENT: Community Medicine, Teerthanker Mahaveer Medical College & Research Centre**

On the behalf of TMU Moradabad Institutional Ethics Committee (TMU-IEC), I hereby communicate to you that ethical approval has been granted for your research project protocol by the committee on **21-11-2023** with respect of the undertakings mentioned in the above project. Should any other methodology be used, these require separate authorization.

The investigator may therefore commence with the research as from the date of this certificate, using the reference number indicated above.

Please note that the TMU IEC must be informed immediately of:

- Any material change in the conditions or undertakings mentioned in the document.
- Any material breaches of ethical undertakings or events that impact upon the ethical conduct of the research.

The Principal Investigator must report to the TMU-IEC in the prescribed format, where applicable, bi-annually, and at the end of the project, in respect of ethical compliance.

TMU-IEC retains the right to withdraw or amend this **ETHICAL CLEARANCE CERTIFICATE** if:

- Any unethical principal or practices are revealed or suspected
- Relevant information has been withheld or misrepresented

TMU-IEC shall have an access to any information or data at any time during the course or after completion of the project.

On behalf of Ethics Committee, I wish you well in your research.

DR. ROHIT VARSHNEY
MEMBER SECRETARY
INSTITUTIONAL ETHICS COMMITTEE
TEERTHANKER MAHAVEER UNIVERSITY
MORADABAD

**Dr. Rohit Varshney**
Member Secretary
University, Institutional Ethics Committee
